# T cell reactivity to the SARS-CoV-2 Omicron variant is preserved in most but not all prior infected and vaccinated individuals

**DOI:** 10.1101/2022.01.04.21268586

**Authors:** Vivek Naranbhai, Anusha Nathan, Clarety Kaseke, Cristhian Berrios, Ashok Khatri, Shawn Choi, Matthew A. Getz, Rhoda Tano-Menka, Onosereme Ofoman, Alton Gayton, Fernando Senjobe, Kerri J. St Denis, Evan C. Lam, Wilfredo F. Garcia-Beltran, Alejandro B. Balazs, Bruce D. Walker, A. John Iafrate, Gaurav D. Gaiha

## Abstract

The SARS-CoV-2 Omicron variant (B.1.1.529) contains mutations that mediate escape from infection and vaccine-induced antibody responses, although the extent to which these substitutions in spike and non-spike proteins affect T cell recognition is unknown. Here we show that T cell responses in individuals with prior infection, vaccination, both prior infection and vaccination, and boosted vaccination are largely preserved to Omicron spike and non-spike proteins. However, we also identify a subset of individuals (∼21%) with a >50% reduction in T cell reactivity to the Omicron spike. Evaluation of functional CD4^+^ and CD8^+^ memory T cell responses confirmed these findings and reveal that reduced recognition to Omicron spike is primarily observed within the CD8^+^ T cell compartment. Booster vaccination substantially enhanced T cell responses to Omicron spike. In contrast to neutralizing immunity, these findings suggest preservation of T cell responses to the Omicron variant, although with reduced reactivity in some individuals.

## INTRODUCTION

The severe acute respiratory syndrome coronavirus 2 (SARS-CoV-2) Omicron variant (B.1.1.529), first identified in November 2021, has been the cause of a new surge of infections globally (Viana et al., 2021). With as many as 36 substitutions in the viral spike protein and 59 mutations in total throughout its genome, Omicron has been found to evade neutralization by infection- and vaccine-induced antibodies with unprecedented frequency (Garcia-Beltran et al., 2021a; Hoffmann et al., 2021) and escape neutralization by most therapeutic monoclonal antibodies (Ikemura et al., 2021; VanBlargan et al., 2021). Additional booster vaccine doses partially compensate for this effect (Garcia-Beltran et al., 2021b; Hoffmann et al., 2021), but the durability of such protective antibody response remains to be determined. Thus, whether additional arms of the adaptive immune response, namely T cell responses, can augment protection against Omicron infection and disease is of considerable interest and has implications for predicting the course of future SARS-CoV-2 variants.

In individuals with previous SARS-CoV-2 infection and vaccinees, robust T cell responses are quantitatively and qualitatively associated with milder outcomes (Rydyznski Moderbacher et al., 2020). Early induction of antigen-specific CD4^+^ T cells following vaccination is associated with coordinated generation of antibody and CD8^+^ T cell responses (Painter et al., 2021). Previous studies have also shown a key role for CD8^+^ T cells in mitigating COVID-19 disease severity and inducing long-term immune protection. Mild COVID-19 disease is associated with increased clonal expansion of CD8^+^ T cells in bronchoalveloar lavage fluid (Liao et al., 2020), robust CD8^+^ T-cell reactivity to SARS-CoV-2 epitopes (Peng et al., 2020; Sekine et al., 2020), and rapid CD8^+^ T cell-mediated viral clearance (Tan et al., 2021). In addition, depletion of CD8^+^ T cells from convalescent macaques reduced protective immunity (McMahan et al., 2020). Given that T cells can target regions across the SARS-CoV-2 proteome and are not limited solely to the spike protein, it is perhaps not unexpected that prior SARS-CoV-2 variants were able to escape neutralizing antibody responses (Garcia-Beltran et al., 2021a) but not T cells (Geers et al., 2021). Thus, in light of the emergence of the Omicron variant, we sought to determine the extent to which mutations in the variant spike and non-spike proteins affect CD4^+^ and CD8^+^ T cell reactivity.

Utilizing samples from prior SARS-CoV-2 infected, vaccinated, and both prior infected and vaccinated individuals, we found that circulating effector T cell responses and both CD4^+^ and CD8^+^ memory T cell responses were generally preserved to the Omicron variant. However, distinct from previous variants of concern (VOC), such as Delta, a subset of individuals had reduced effector and memory T cell recognition to the Omicron spike protein relative to wildtype spike, with a particularly noticeable effect on spike-specific CD8+ T cell memory responses. Booster doses enhanced the magnitude of responses to wildtype and Omicron spike, although did not completely mitigate the comparatively reduced T cell reactivity to Omicron in individual participants. These findings therefore have important implications in ascertaining the role of immune responses in morbidity and mortality due to Omicron and may inform the development variant-specific and variant-resistant second-generation vaccines.

## RESULTS

To assess the cross-reactivity of T cell responses to the Omicron variant, we studied anti-SARS-CoV-2 T cell responses in 76 ambulatory adult volunteers in Chelsea, Massachusetts sampled prior to vaccination, after primary series vaccination, and/or after receipt of additional ‘booster’ doses. Study groups were stratified by prior infection (confirmed by anti-nucleocapsid antibody testing) and vaccination status (Table S1). In total, we studied 101 samples from 76 donors (Figure 1A). The median age was 45 years (range 37–60 years) and 64% were female. Of the previously infected individuals, we included 11 unvaccinated, 12 vaccinated sampled after initial vaccination series, and 13 vaccinated and sampled after booster doses. Among individuals without previous infection, we included 10 unvaccinated, 24 sampled after initial vaccination series, and 31 vaccinated and sampled after booster doses. Samples were obtained at a median 220 (range 130–286) days after primary series vaccination or 10 days (range 8–54) after additional booster doses. The primary analysis of host, vaccine, and variant variables that affect T cell responses was by multivariate regression.

**Figure 1.**
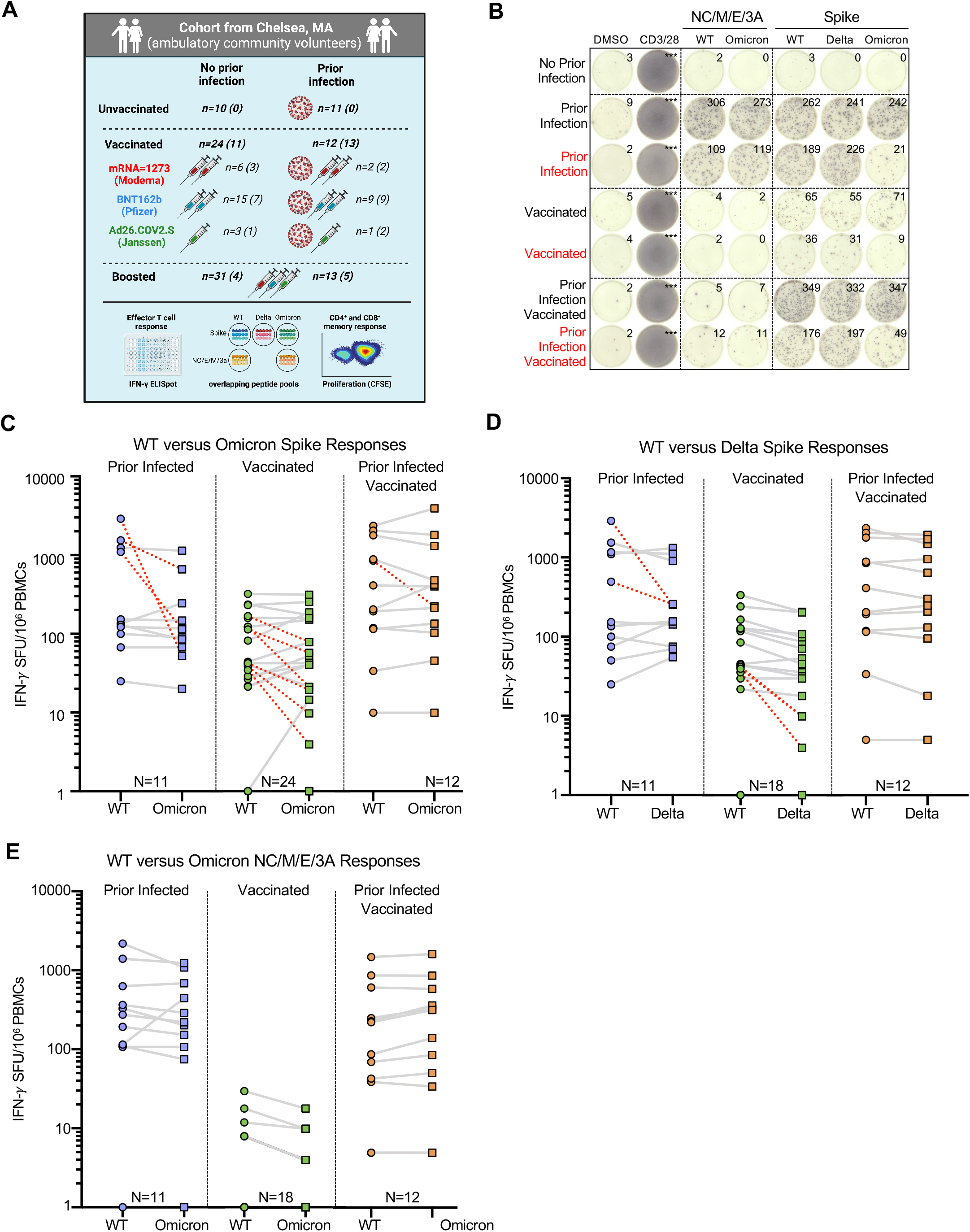
Effector T cell reactivity to the SARS-CoV-2 Omicron variant is preserved in most but not all individuals with prior infection and/or primary series vaccination. **(A)** Schematic of the study created with Biorender.com: Participants were enrolled in Chelsea, Massachusetts and stratified according to whether they had documented asymptomatic or symptomatic SARS-CoV-2 infection (ascertained by anti-nucleocapsid antibody testing) and vaccination status (see Table S1). 101 PBMC samples from 76 individuals were studied. 25 individuals provided samples prior to and after receipt of additional booster vaccine doses. Total (CD4^+^ and CD8^+^) effector T cell reactivity to SARS-CoV-2 overlapping peptide pools from wildtype, Omicron or Delta spike and from wildtype or Omicron non-spike structural and accessory proteins (nucleocapsid/membrane/envelope/ORF3A, i.e. NC/M/E/3A) was assessed by IFN-g ELISpot (the number is shown for each group). CD4^+^ and CD8^+^ memory T cell response to wildtype or Omicron spike was assessed in a subset of participants by CFSE-based proliferation assay (see Figure 3). Numbers for each group are shown in parentheses. Table S2 describes the degree of overlap in peptide pools. **(B)** Representative IFN-γ ELISpot responses for study participants following no stimulation (dimethyl sulfoxide DMSO only), anti-CD3 and anti-CD28 stimulation (positive control), overlapping NC/M/E/3A peptide pools from wildtype SARS-CoV-2 and Omicron variant and overlapping spike peptide pools from wildtype, Delta and Omicron variant. Those delineated in red are representative examples of individuals with >50% decreased T cell responses to the Omicron spike peptide pool compared to wildtype. **(C)** Comparative IFN-γ ELISpot spot forming units (SFUs) per 10^6^ peripheral blood mononuclear cells (PBMCs) in individuals with prior infection, vaccination and both prior infection and vaccination to overlapping wildtype and Omicron spike peptide pools. Overall T cell responses to wildtype and Omicron were comparable across all groups by multivariate regression, although red dashed lines indicate the 10 participants with a >50% decrease (0.3log_10_) in reactivity. **(D)** Comparative IFN-γ ELISpot responses in individuals with prior infection, vaccination and both prior infection and vaccination to overlapping wildtype and Delta spike peptide pools. Overall T-cell responses to wildtype and Delta were comparable across all groups, although red dashed lines indicate the 4 participants with a >50% decrease (0.3log_10_) in reactivity. (**E**) Comparative IFN-γ ELISpot responses in individuals with prior infection, initial vaccination series and both prior infection and vaccination to overlapping peptide pools of the wildtype and Omicron NC/M/E/3A. In C-E, each dot is a single participant. Circles denote responses to wildtype peptides and squares to Omicron or Delta peptides. Dots are colored by prior infection and vaccine stratum (blue for prior infection and no vaccination, green for no prior infection and vaccination with primary series and orange for both prior infection and vaccination with primary series). In C-E, pairwise comparison of effector T cell reactivity towards wildtype versus variant by a paired t-test (not adjusted for multiple comparisons or covariates) was not significant.

### Effector T cell reactivity to the SARS-CoV-2 Omicron variant is preserved in most but not all prior infected and vaccinated individuals

To assess the total (CD4^+^ and CD8^+^) effector T cell response, we performed an IFN-γ ELISpot following stimulation with pooled overlapping 15mer peptides spanning the full length of the wildtype, Delta (B.1.617.2), or Omicron (B.1.1.529) spike protein and the non-spike SARS-CoV-2 structural and accessory proteins (nucleocapsid, membrane, enveloped and open reading frame 3A, i.e. NC/M/E/3A) from wildtype and Omicron. The evaluated peptides span the full-length of spike: relative to wildtype, 27.3% (86/315) spike peptides were unique to Omicron and 8.6% (27/315) to Delta. For the NC/M/E/3A pools, the evaluated peptides span the full-length of the respective proteins: relative to wildtype, 10.1% (24/237) of the peptides were unique to Omicron (Table S2). In the primary multivariate analysis of T cell reactivity (Table S3), the magnitude of effector T cell responses to spike and non-spike proteins did not vary by variant, and was not affected by age, sex and primary vaccine series. However, examination of individual responses reveals specific patients in whom responses to the Omicron spike were reduced by >50% (Figure 1B, denoted in red). Prior infection, duration after primary series vaccination, and receipt of an additional ‘booster’ dose were independently associated with magnitude of response (Table S3).

The median effector T cell reactivity against wildtype and Omicron spike (in SFU/10^6^ PBMC) was 152 and 114 for individuals with prior infection; 43 and 42 for individuals after primary series vaccination (without prior infection); and 311 and 315 for individuals with prior infection after primary series vaccination (Figure 1C). In comparison, the median effector T cell reactivity against delta spike (in SFU/10^6^ PBMC) was 155 for individuals with prior infection; 34 for individuals after primary series vaccination (without prior infection); and 277 for individuals with prior infection after primary series vaccination (Figure 1D). Regardless of variant, prior infection was associated with a higher magnitude of effector T cell responses (0.55 log_10_ SFU/10^6^ PBMC higher response, 95%CI 0.38-0.72, p<0.001). Effector T cell responses declined modestly over time (−0.02 log_10_ SFU/10^6^ PBMC lower response per week, 95%CI –0.05,0.00, p=0.028). Neither age nor sex influenced responses, and in this analysis primary vaccine type was not associated with differences in responses (Table S3). Surprisingly, 21.2% (10/47) of participants with prior infection and/or vaccination had a >50% (0.3log_10_) reduction in T cell response to Omicron spike (denoted in red in Figure 1C), with 12.7% of participants (6/47) having a >70% (0.5 log_10_) reduction. In contrast, only 9.7% (4/41) of participants with prior infection and/or vaccination had a >50% (0.3log_10_) reduction in overall effector T cell response to Delta spike. Thus, while T cell responses induced by prior infection and/or vaccination are broadly cross-reactive at a population level, a distinct subset of individuals have substantially reduced T cell recognition of the mutated Omicron spike protein.

In contrast to spike-specific T cell responses, T cell reactivity against wildtype and Omicron NC/M/E/3A was preserved in all individuals with prior infection (with or without subsequent vaccination). The median effector T cell reactivity against wildtype and Omicron NC/M/E/3A (in SFU/10^6^ PBMC) was 275 and 220 for individuals with prior infection; 1 and 0 for individuals after primary series vaccination (without prior infection); and 160 and 237 for individuals with prior infection and after primary series vaccination (Figure 1E). In individuals with prior infection or prior infection and vaccination, the magnitude of reactivity towards NC/M/E/3A was correlated with that of spike for wildtype and Omicron peptides (Figure S1).

### Additional booster vaccine doses enhance effector T cell responses to SARS-CoV-2 wildtype and omicron

Individuals with prior infection demonstrated higher T cell responses to spike, suggesting that repeated exposure to antigen may potentially enhance cross-reactive T cell responses. We therefore assessed the impact of booster vaccination on T cell reactivity by IFN-γ ELISpot. Similar to the evaluation of pre-boost samples, overall effector T cell responses towards wildtype and Omicron spike across our study participants were comparable post-booster (Figure 2B). Moreover, receipt of a booster dose was associated with a 1.1log_10_ SFU/10^6^ PBMC increase (95% CI 0.91-1.2, p<0.001) in the magnitude of T cell response (Table S3), with specific fold increases of 20.1 against wildtype and 20.4 against Omicron in 25 participants with paired sampling (Figure 2C). However, even after booster vaccination, 9.1% (4/44) participants still demonstrated >50% reduced reactivity to Omicron spike relative to wildtype. Overall, the frequency of >50% reduced effector T cell responses to the Omicron variant was more frequent than to Delta in 85 individual sample points with both measures (Fisher’s exact p-value 0.023, Table S4).

**Figure 2.**
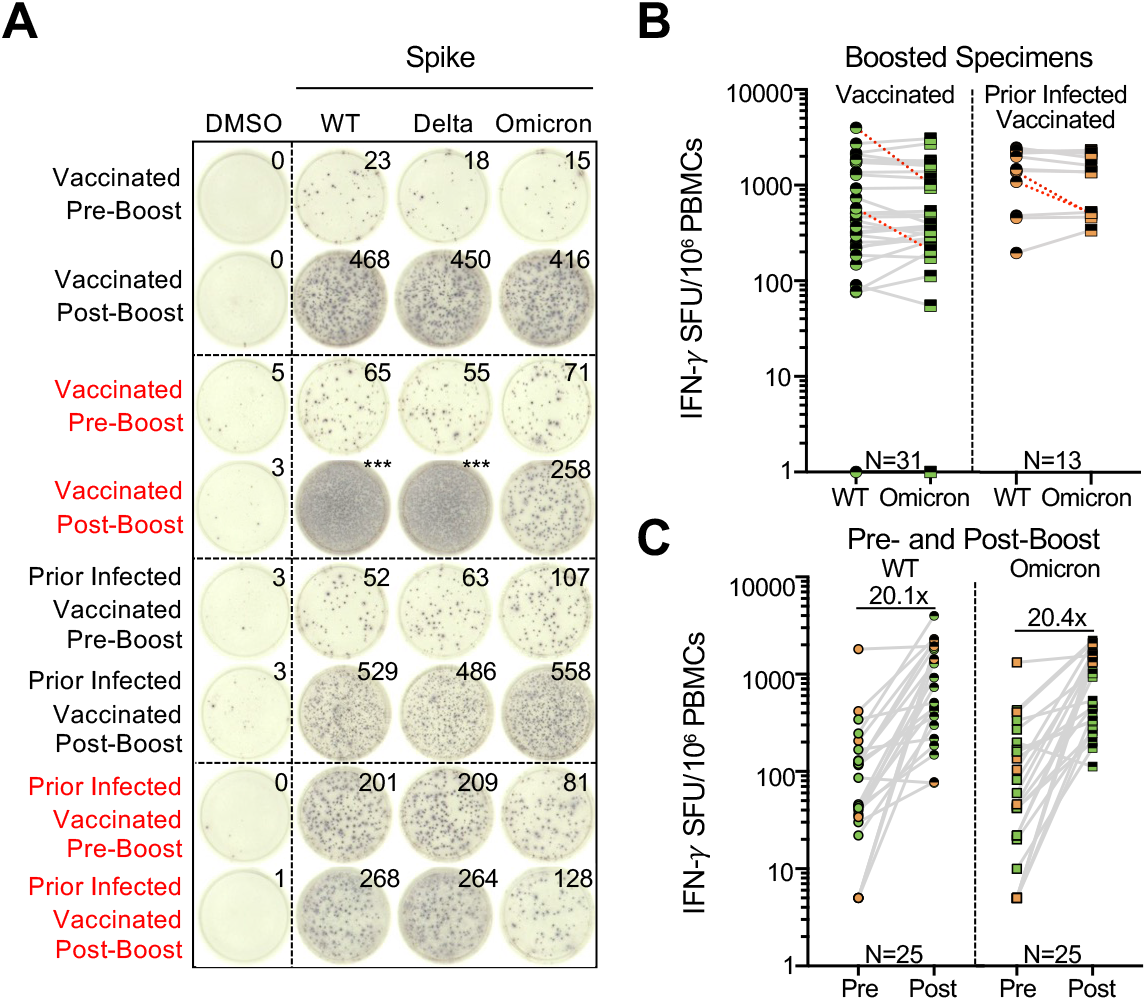
Effector T cell responses to SARS-CoV-2 wildtype and Omicron are enhanced by additional booster vaccination. **(A)** Representative IFN-γ ELISpot responses for study participants. Shown are IFN-γ ELISpot responses following no stimulation (dimethyl sulfoxide DMSO only), anti-CD3 and anti-CD28 stimulation (positive control), and overlapping spike protein peptide pools from wildtype and Omicron variant in individuals with no prior infection and vaccinated or with prior infection and vaccinated sampled prior to receipt of booster (pre-boost) or after booster (post-boost) vaccine doses. Those delineated in red indicate representative examples of individuals with >50% decreased T cell responses to the Omicron spike peptide pool in comparison with wildtype. **(B)** Comparative IFN-γ ELISpot responses in individuals with and without prior infection following booster vaccination (range 8-54 days following booster dose). Red dashed lines indicate the 4 participants with a >50% decrease (0.3log_10_) in T cell reactivity to Omicron relative to wildtype. **(C)** Comparative T cell reactivity pre- and post-booster vaccination (8-10 days following booster dose) in 25 participants to both wildtype and Omicron spike protein. Booster vaccination elicited an approximately 20-fold increase in T cell response magnitude to both Spike proteins. In B and C, each dot is a single participant. Circles denote responses to wildtype peptides and squares to Omicron peptides. Dots are colored by prior infection and vaccine stratum (green for no prior infection and vaccination with primary series and orange for both prior infection and vaccination with primary series). Fill denotes sampling pre-boost (full filled) or post-boost (half filled). In B, pairwise comparison of effector T cell reactivity towards wildtype versus variant by a paired t-test (not adjusted for multiple comparisons or covariates) was not significant.

### Proliferative CD4+ memory T cell responses are preserved against Omicron but CD8^+^ T cell responses are reduced

To assess the cross-reactivity of functional CD4^+^ and CD8^+^ memory T cell responses to Omicron, we performed a 6-day carboxyfluorescein succinimidyl ester (CFSE) proliferation assay (Figure S2) on samples from individuals who were vaccinated and/or previously infected and/or received booster vaccine doses (n = 33 participants) using overlapping wildtype or Omicron spike peptide pools. We felt it was important to utilize this assay given that antigen-specific proliferation has been strongly associated with functional T cell responses and cytotoxicity (Migueles et al., 2002, 2008; Ndhlovu et al., 2013). The patient cohort studied here was a subset of that used for the IFN-γ ELISpot assay, wherein 11 samples were from vaccinated individuals, 13 from previously infected and vaccinated individuals, and nine from vaccinated and boosted individuals (five of whom were also previously infected). A paired t-test demonstrated that while the magnitude of proliferative spike-specific CD4^+^ T cell responses did not vary by variant, proliferative CD8+ T cell responses to Omicron spike were decreased compared to wildtype in previously infected, vaccinated participants (p = 0.009) and across all study participants (p < 0.005), which is further illustrated by examination of individual patient responses (Figure 3A, denoted in red). CD4^+^ proliferative responses remained largely cross-reactive to Omicron spike, with only 12% (4/33) of individuals with prior infection and/or vaccination and/or booster showing a >50% (0.3log_10_) reduction (Figure 3B). A larger proportion of 39% of individuals (13/33) exhibited a decreased CD8^+^ T cell proliferative response to Omicron spike (Figure 3C). A multivariate regression analysis revealed that neither age nor sex influenced CD4^+^ or CD8^+^ T cell responses but proliferative CD8^+^ responses tended to be lower for Omicron vs wildtype after adjusting for all covariates and were significantly increased by booster doses (Table S5). These data indicate that the reduced reactivity in a subset of individuals to the Omicron spike protein is primarily observed in the CD8^+^ T cell compartment, although this can be enhanced with booster vaccination.

**Figure 3:**
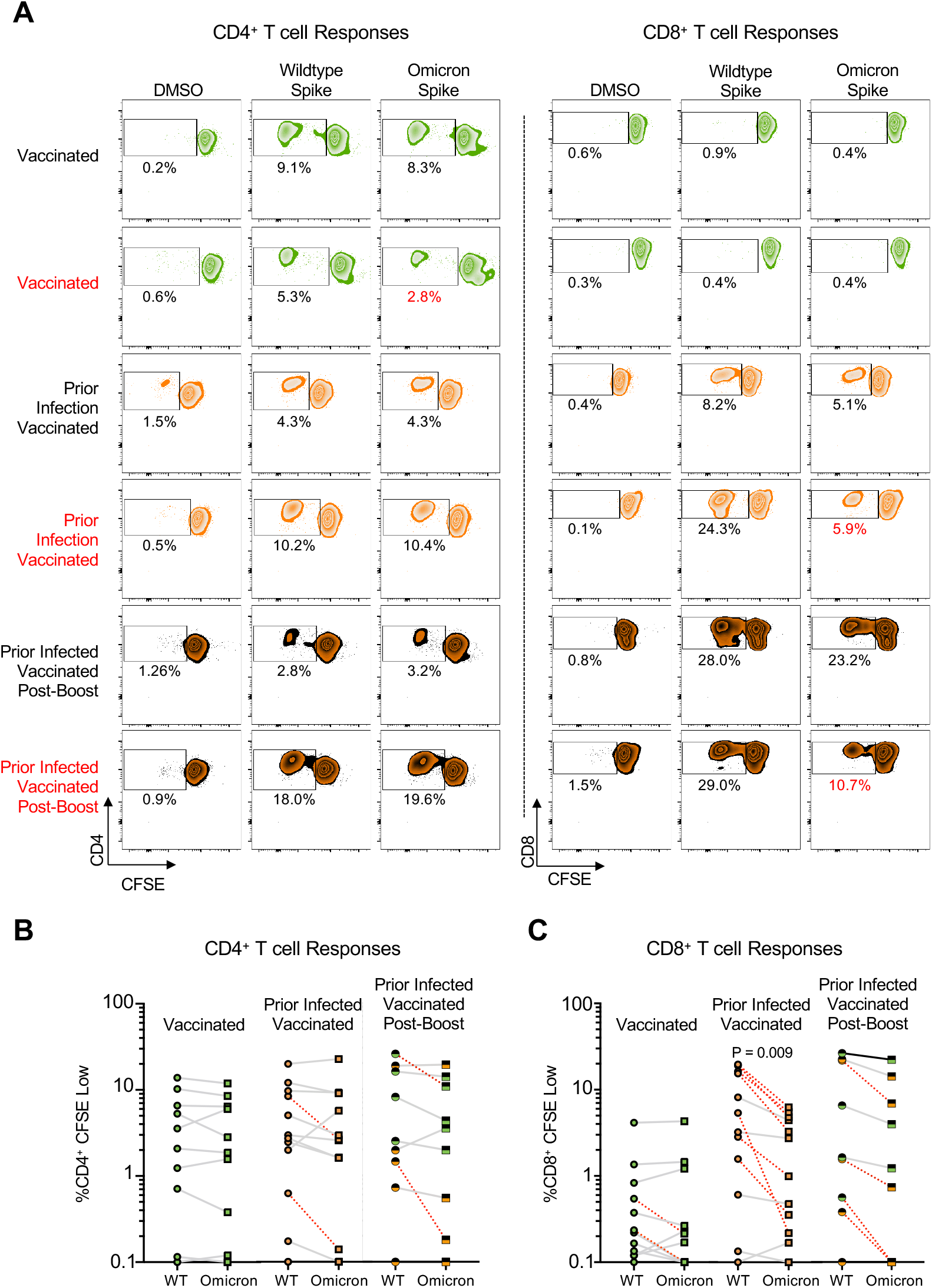
Proliferative spike-specific CD4^+^ T cell responses are preserved against Omicron but CD8^+^ responses are reduced. **(A)** Representative CFSE responses for study participants. Shown are CD4^+^ (left) and CD8^+^ (right) responses following no stimulation (dimethyl sulfoxide DMSO only) and overlapping spike protein peptide pools from wildtype and Omicron variant. Those delineated in red indicate representative examples of individuals with >50% decreased T-cell responses to the Omicron spike peptide pool in comparison with wildtype. **(B)** Comparative %CD4^+^ CFSE Low cells in individuals with vaccination, both prior infection and vaccination, and prior infection, initial vaccination, and booster vaccination in response to overlapping wildtype and Omicron spike peptide pools. Red dashed lines indicate the 4 individuals with a >50% decrease (0.3log_10_) in proliferative response. Pairwise comparison of memory CD4^+^ T-cell proliferation to wildtype vs. Omicron spike by a paired t-test (not adjusted for multiple comparisons or covariates) was not significant. **(C)** Comparative %CD8^+^ CFSE Low cells in individuals with vaccination, both prior infection and vaccination, and prior infection, initial vaccination, and booster vaccination in response to overlapping wildtype and Omicron spike peptide pools. Red dashed lines indicate the 13 individuals with a >50% decrease (0.3log_10_) in proliferative response. Pairwise comparison of proliferative CD8^+^ T cell responses in previously infected, vaccinated individuals (p = 0.009) and across all study participants (p < 0.005) to wildtype vs. Omicron spike by a paired t-test (not adjusted for multiple comparisons or covariates) revealed a significant reduction in response to Omicron spike.

### Effector T cell responses to Omicron are preserved even in individuals with undetectable neutralization of Omicron

Within this cohort of individuals, we recently reported markedly reduced neutralization of Omicron following primary series vaccination, which was overcome by additional ‘booster’ doses (Garcia-Beltran et al., 2021b). Neutralization and T cell responses against wildtype SARS-CoV-2 were correlated in magnitude in the subset of individuals with overlapping measures (Figure 4). Although additional booster doses increased both antibody and effector T cell responses, many individuals who had undetectable neutralization of Omicron pseudotyped virus had measurable T cell responses against Omicron spike prior to receipt of a booster dose. Using a pseudoneutralization titer threshold of 20 (Garcia-Beltran et al., 2021c) and a T cell response threshold of 23.3 SFU/10^6^ PBMC (the maximal response detected among unvaccinated individuals without prior infection), 4/4 prior infected vaccinated individuals and 8/15 (no prior infection) vaccinated individuals who had low neutralization had measurable T cell responses. However, 38.9% (7/18) of individuals vaccinated with the primary series without prior infection demonstrated T cell reactivity and neutralization of the Omicron variant beneath the above-described threshold.

**Figure 4:**
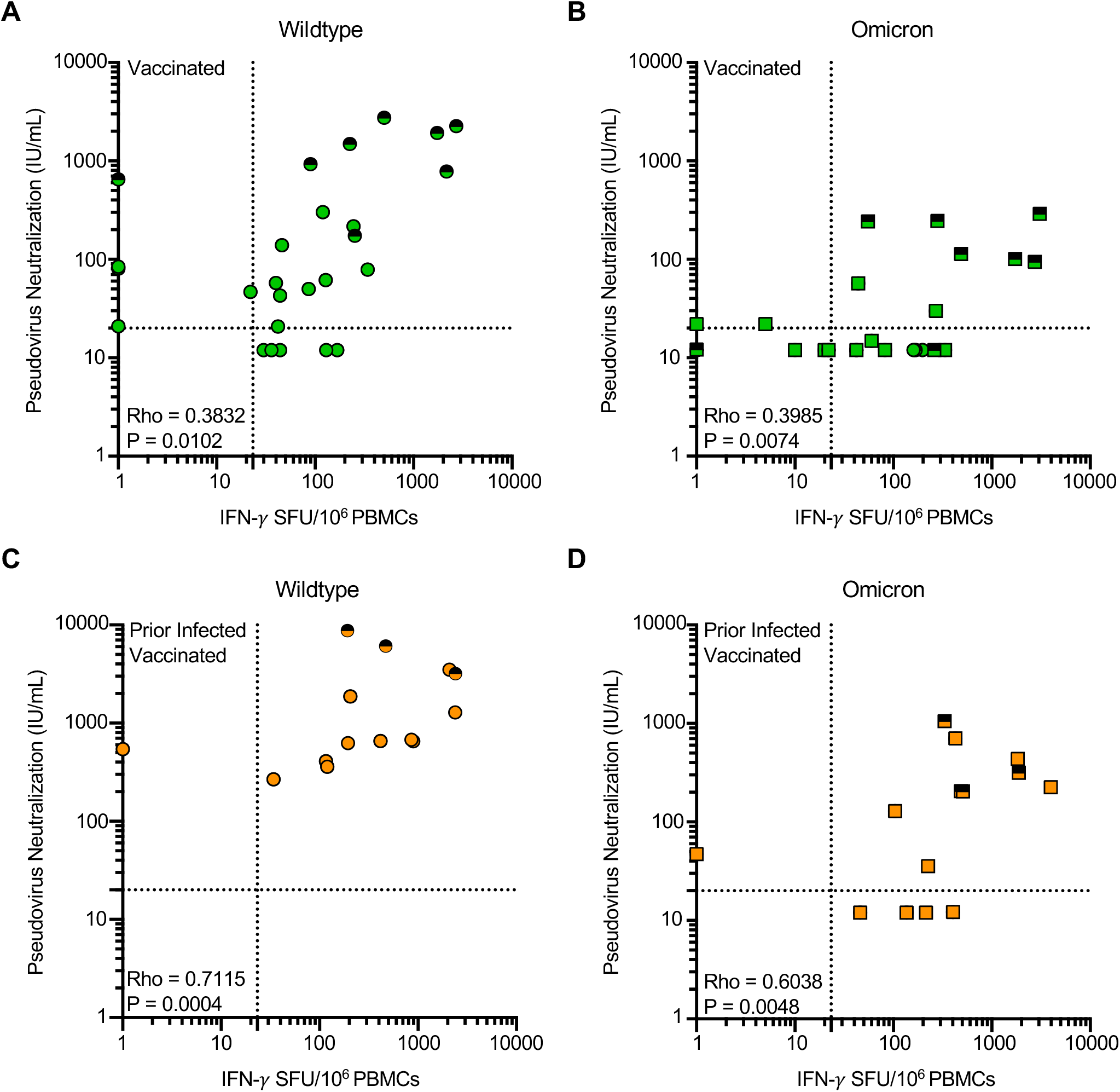
Effector T cell responses to omicron are present in individuals with undetectable neutralization of omicron. Shown are the correlations between magnitude of effector T-cell response (IFN-γ SFU per 10^6^ PBMCs) and pseudovirus neutralization (IU/mL) against wildtype (Panel A and C) or Omicron (B and D) spike. Serum neutralization of pseudotyped virus entry into ACE2-expressing 293T cells was previously reported in the same participants at the same timepoints (Garcia-Beltran et al., 2021b). In A-D each dot is a single participant. Circles denote responses to wildtype peptides and squares to Omicron peptides. Dots are colored by prior infection and vaccine stratum (green for no prior infection and vaccination with primary series and orange for both prior infection and vaccination with primary series). Fill denotes sampling pre-boost (full filled) or post-boost (half-filled). Spearman correlation coefficients are denoted in each panel. Dotted lines denote a pseudoneutralization titer threshold of 20 (Garcia-Beltran et al., 2021c) and a T cell response threshold of 23.3 SFU/10^6^ PBMC (the maximal response detected among unvaccinated individuals without prior infection).

## DISCUSSION

Immune responses induced by SARS-CoV-2 infection and/or vaccination induce a composite of antibody, effector T cell and memory T cell responses that target viral antigens subject to mutation. In this report, we evaluated whether existing anti-SARS-CoV-2 T cell responses are cross-reactive towards the Omicron variant or differ in comparison to wildtype and the Delta variant. We found that, in aggregate, the magnitude of circulating effector T cell responses towards Omicron spike and non-spike structural proteins were conserved across variants and enhanced by additional booster vaccine doses. However, examination of individual responses revealed that a distinct proportion of individuals with prior infection and/or vaccination have substantially reduced T cell reactivity to Omicron (but not Delta). Further evaluation of the spike-specific CD4+ and CD8+ memory T cell compartment revealed a significant difference in CD8+ T cell proliferation in response to Omicron spike relative to wildtype. These findings are consistent with the modestly higher degree of antigen diversity in Omicron, although remain somewhat unexpected given that the vast majority of the spike protein remains largely sequence conserved (97.2%, i.e. 1237/1273 amino acids unchanged). Therefore, in some individuals, it is possible that Omicron variation may mediate escape from specific HLA-restricted T cell responses induced by prior infection and vaccination. In sum, T cell reactivity to the SARS-CoV-2 Omicron variant was preserved in most but not all prior infected and vaccinated individuals.

The SARS-CoV-2 Omicron variant demonstrates substantial escape from neutralizing antibody responses (Garcia-Beltran et al., 2021b; Hoffmann et al., 2021) likely due to the striking enrichment of mutations at key sites in the receptor binding domain (RBD) that are critical for neutralization by antibodies (Greaney et al., 2021). In contrast, we found that T cell reactivity was relatively preserved in most individuals against Omicron, and in many individuals with undetectable omicron neutralizing antibody responses, effector T cell responses were measurable. Previous studies have identified an association between T cell immunity and mild COVID-19 disease (Peng et al., 2020; Rydyznski Moderbacher et al., 2020; Tan et al., 2021). In addition, in animal models of SARS-CoV-2 (McMahan et al., 2020), T cell responses appear to be important in reducing disease acquisition and severity. Thus, the high frequency of preserved T cell responses against Omicron suggests that T cell responses may be responsible for vaccine effectiveness (and also from natural infection) against severe outcomes from Omicron infection that appear higher than predicted by absent or lower neutralization.

Prior SARS-CoV-2 infection, despite being remote, was associated with higher T cell effector and memory responses than vaccination alone, and responses were directed against both spike and non-spike proteins in contrast to being focused solely on spike. This may reflect the impact of distinct antigen kinetics and multiple antigen exposures during infection leading to qualitatively different responses in comparison to vaccination. The preservation of T cell reactivity to non-spike structural and accessory proteins in all individuals is likely due to the substantially reduced number of mutations within NC/M/E/3A relative to spike, suggesting that these proteins may be highly attractive for second-generation COVID-19 vaccines. In particular, vaccine strategies that induce robust memory and effector T cell responses alongside antibody responses which are collectively targeted against conserver, variant-resistant sites (Meyers et al., 2021; Nathan et al., 2021) may yield more durable T cell immunity capable of providing broad protection against future variants.

Collectively, these data provide insight into the immune mechanisms that may account for clinical observation of Omicron pathophysiology and demonstrate the contribution of vaccine boosters to enhancing cellular immunity to SARS-CoV-2 variants. These findings also support continued evaluation of second-generation vaccine approaches that induce robust T cell responses that target both variant spike and non-spike antigens in order to overcome current and future SARS-CoV-2 evolution.

## LIMITATIONS OF STUDY

Our study has some noteworthy limitations. First, this is a study of dynamic immune responses which have distinct kinetics but timing of sampling was constrained to ∼6 months after primary series vaccination and sooner after booster doses. Secondly, although we included individuals with prior infection, primary series vaccination and booster vaccines with the three vaccines deployed in the USA (mRNA1273, BNT162b2 and Ad26.COV2.S), these groups cannot comprehensively capture the variables that may plausibly affect reactivity such as the variant with which individuals were infected, the wide variety of vaccines deployed globally, and the differences in the timing of additional doses. Thirdly, we employed IFN-γ ELISPOT and proliferation assays to estimate T cell responses. While these assays are highly sensitive for functionally relevant T cell responses, additional assays, such as the activation induced marker assay (Grifoni et al., 2020) or intracellular cytokine staining following peptide stimulation could also be utilized.

## Data Availability

All data produced in the present work are contained in the manuscript

## ACKNOWLEDGEMENTS, FUNDING SUPPORT

We thank Shiv Pillai, MD PhD for excellent advice on this manuscript. We thank Anand Dighe, MD, Andrea Nixon, BS, and the MGH Core Laboratory for excellent assistance with clinical SARS-CoV-2 serology testing. This work was supported by the Peter and Ann Lambertus Family Foundation. This study was supported by NIH grants P01 DK011794-51A1 (A.K.), R01AI149704 (B.D.W.), UM1AI144462 (G.D.G. and B.D.W.), and DP2AI154421 (G.D.G.) and a grant from the MassCPR (B.D.W. and G.D.G.). Additional support was provided by the Howard Hughes Medical Institute (B.D.W.), the Ragon Institute of MGH, MIT and Harvard (B.D.W. and G.D.G.), the Mark and Lisa Schwartz Foundation and Enid Schwartz (B.D.W.), and Sandy and Paul Edgerly. V.N. received support from a Medscape Young Investigators Lung Cancer award. A.B.B. was supported by the National Institutes for Drug Abuse (NIDA) Avenir New Innovator Award DP2DA040254, the MGH Transformative Scholars Program and a Massachusetts Consortium on Pathogenesis Readiness (MassCPR) grant. G.D.G. is supported by the Bill and Melinda Gates Foundation, a Burroughs Wellcome Career Award for Medical Scientists and the Gilead HIV Research Scholars Program.

## AUTHOR CONTRIBUTIONS

Conceptualization, V.N., B.D.W., A.J.I., G.D.G.; Formal Analysis, V.N., A.N., G.G.; Methodology and Investigation, V.N., A.N., C.K., C.B., M.A.G., R.TM., O.O., A.G., F.S., K.J.SD., E.C.L., W.F.G.B., A.B.B., G.D.G.; Resources, A.K., S.C.; Writing – Original Draft, V.N., A.N., G.G.; Funding Acquisition, A.B.B, B.D.W., A.J.I., G.G.; Supervision, A.B.B., A.J.I., G.D.G.

## INCLUSION AND DIVERSITY

We worked to ensure gender balance in the recruitment of human subjects. We worked to ensure ethnic or other types of diversity in the recruitment of human subjects. We worked to ensure that the study questionnaires were prepared in an inclusive way. One or more of the authors of this paper self-identifies as an underrepresented ethnic minority in science. One or more of the authors of this paper self-identifies as a member of the LGBTQ+ community.

## STAR ⍰ METHODS

### KEY RESOURCES TABLE

| REAGENT or RESOURCE | SOURCE | IDENTIFIER |
| --- | --- | --- |
| **Antibodies** |  |  |
| Mouse monoclonal anti-human CD3 (clone OKT3) | Biolegend | Cat# 317302;<br>RRID:AB_571927 |
| Mouse monoclonal anti-human CD28 (clone CD28.2) | Biolegend | Cat# 302902;<br>RRID:AB_314304 |
| Mouse monoclonal anti-human CD3 (clone SK7) labeled with PE-Cy7 fluorophore | Biolegend | Cat# 344816;<br>RRID:AB_10640737 |
| Mouse monoclonal anti-human CD4 (clone RPA-T4) labeled with BV711 fluorophore | Biolegend | Cat# 300558; |
| Mouse monoclonal anti-human CD8 (clone SK1) labeled with APC fluorophore | Biolegend | Cat# 980904;<br>RRID:AB_2564393 |
| LIVE/DEAD Violet Viability | Life Technologies | Cat# L34960 |
| **Bacterial and virus strains** |  |  |
| **Biological samples** |  |  |
| PBMC Specimens of prior COVID infected, vaccinated, both infected and vaccinated and booster vaccinated individuals | MGH | N/A |
| **Chemicals, peptides, and recombinant proteins** |  |  |
| PepMix SARS-CoV-2 Wildtype Spike Peptide Pool | JPT | PM-PM-WCPV-S-1 |
| PepMix SARS-CoV-2 Wildtype Nucleoprotein Peptide Pool | JPT | PM-WCPV-NCAP-1 |
| PepMix SARS-CoV-2 Wildtype Membrane Peptide Pool | JPT | PM-WCPV-VME-1 |
| PepMix SARS-CoV-2 Wildtype Envelope Peptide Pool | JPT | PM-WCPV-VEMP-1 |
| PepMix SARS-CoV-2 Protein 3a Peptide Pool | JPT | PM-WCPV-AP3A-1 |
| PepMix SARS-CoV-2 B.1.617.2 (Delta) Spike Peptide Pool | JPT | PM-SARS2-SMUT06-1 |
| SARS-CoV-2 B.1.1.529 (Omicron) Spike Peptide Pool | MGH Peptide Core | N/A |
| CellTrace CFSE Cell Proliferation | Life Technologies | Cat# C34554 |
| **Critical commercial assays** |  |  |
| Human IFN-gamma ELISpot Basic Kit | MABTECH | Cat# 3420-2A |
| **Deposited data** |  |  |
| **Experimental models: Cell lines** |  |  |
| **Experimental models: Organisms/strains** |  |  |
| **Oligonucleotides** |  |  |
| **Recombinant DNA** |  |  |
| **Software and algorithms** |  |  |
| **Other** |  |  |

## RESOURCE AVAILABILITY

### Lead Contact

Further information and requests for resources and reagents should be directed to and will be fulfilled by the lead contact, Gaurav D. Gaiha.

### Materials Availability

All requests for resources and reagents should be directed to and will be fulfilled by the lead contact. All reagents will be made available on request after completion of a Materials Transfer Agreement.

### Data and Code Availability

All data supporting the findings of this study available within the paper and available from the lead contact upon request.

## EXPERIMENTAL MODEL AND SUBJECT DETAILS

### Human subjects

Use of human samples was approved by Massachusetts General Brigham Institutional Review Board (protocol 2020P001081). Consenting ambulatory adults in Chelsea, Massachusetts were enrolled in a study of immune responses and sampled in mid-2020 or December 2021. Demographic data, information regarding prior SARS-CoV-2 testing, symptoms, and exposure was collected as was vaccine related information. Prior infection was defined by positive anti-nucleocapsid antibody testing on the Roche Elecsys SARS-CoV-2 assay performed at the MGH clinical laboratory and absence of prior positive SARS-CoV-2 PCR testing. Rates of infection in the Chelsea community were high during early SARS-CoV-2 waves (Naranbhai et al., 2021) and most participants in this study had been infected in the initial waves of infection. The duration from receipt of the final dose of the primary series (first Ad26.COV2.S, or second BNT-162b2 or mRNA-1273) and duration post booster dose were collected and included as covariates as was age and sex. Samples from unvaccinated participants were obtained in 2020 (pre-Omicron period) and the remaining samples of vaccinated, prior infected and vaccinated and boosted individuals were obtained between December 3, 2021 and December 13, 2021. In total, we include 101 samples from 76 individuals; 25 individuals provided pre-booster and post-booster samples.

## METHOD DETAILS

### PBMC Isolation

Blood was collected in heparin tubes and processed within 4 hours of collection. Peripheral blood mononuclear cells (PBMC) were isolated by density gradient sedimentation using Lymphocyte Separation Media (Corning) as per the manufacturer’s instructions and cryopreserved in freezing media consisting of heat-inactivated fetal bovine serum (FBS, Sigma-Aldrich) containing 10% DMSO and stored in liquid nitrogen until use.

### Peptide synthesis and analysis

Complete overlapping 15mer Spike, Nucleocapsid, Membrane, and Envelope peptides (15mer peptide overlapping by 11 amino acids) from the SARS-CoV-2 Omicron (B.1.1.529) variant were synthesized on an automated robotic peptide synthesizers (AAPPTEC, 396 Models MBS, Omega and Apex) by using Fmoc solid-phase chemistry (Behrendt et al., 2016) on 2-chlorotrityl chloride resin (Chatzi et al., 1991). The C-terminal amino acids were loaded using the respective Fmoc-Amino Acids in the presence of DIEA. Unreacted sites on the resin were blocked using methanol, DIEA and DCM (15:5:80 v/v). Subsequent amino acids were coupled using optimized (to generate peptides containing more than 90% of the desired full-length peptides) cycles consisting of Fmoc removal (deprotection) with 25% Piperidine in NMP followed by coupling of Fmoc-AAs using HCTU/NMM activation. Each deprotection or coupling was followed by several washes of the resin with DMF to remove excess reagents. After the peptides were assembled and the final Fmoc group removed, peptide resin was then washed with dimethylformamide, dichloromethane, and methanol three times each and air dried. Peptides were cleaved from the solid support and deprotected using odor free cocktail (TFA/triisopropyl silane/water/DODT; 94/2.5/2.5/1.0 v/v) for 2.5h at room temperature (Teixeira et al., 2002). Peptides were precipitated using cold methyl tertiary butyl ether (MTBE). The precipitate was washed 2 times in MTBE, dissolved in a solvent (0.1% trifluoroacetic acid in 30%Acetonitrile/70%water) followed by freeze drying. Peptides were characterized by Ultra Performance Liquid Chromatography (UPLC) and Matrix Assisted Laser Desorption/Ionization Mass Spectrometry (MALDI-MS). All peptides were dissolved initially in 100% DMSO at a concentration of 40 mM, prior to dilution at the appropriate concentration to create protein-specific peptide pools in RPMI-1640 medium.

### SARS-CoV-2 Antigens

Peptide pools of 15mer sequences (overlapping by 11 amino acids) covering the full length of wildtype spike, nucleocapsid, membrane, envelope and ORF3A were obtained from a commercial source (JPT peptide technologies). Overlapping Delta spike peptide pools were also obtained from JPT. For Omicron spike, nucleocapsid, membrane and envelope peptide pools, 15mer peptides (overlapping by 11 amino acids) covering the full length of the mutated proteins were individually synthesized as crude material (MGH Peptide Core). All peptides were individually resuspended in dimethyl sulfoxide (DMSO) at a concentration of 40 mg/mL. Peptide pools for Omicron spike and non-spike proteins were created by pooling aliquots of individual peptides and resuspension in RPMI and DMSO at 20 µg/mL. Pools were used at a final concentration of 0.25-0.5 µg/mL with an equimolar DMSO concentration in the non-stimulated control.

### *Ex vivo* IFN-γ ELISPOT

IFN-γ ELISpot assays were performed according to the manufacturer’s instructions (Mabtech). PBMCs (1-2×10^5^/well) were incubated with SARS-CoV-2 peptide pools at a final concentration of 0.5□μg/□ml for 16–18h. Anti-CD3 (Clone OKT3, Biolegend, 1ug/mL) and anti-CD28 Ab (Clone CD28.2, Biolegend, 1ug/mL) were used as positive controls. To quantify antigen-specific responses, mean spots of the DMSO control wells were subtracted from the positive wells, and the results were expressed as spot-forming units (SFU) per 10^6^ PBMCs. Responses were considered positive if the results were >5□SFU/10^6^ PBMCs following control subtraction. If negative DMSO control wells had >30 SFU/10^6^ PBMCs or if positive control wells (anti-CD3/anti-CD28 stimulation) were negative, the results were excluded from further analysis. For graphical analyses, negative responses are plotted at a value of 1 SFU/10^6^ PBMCs.

### CFSE Proliferation Assay

PBMCs were suspended at 1 × 10^6^/mL in PBS and incubated at 37°C for 20 min with 0.5 uM carboxyfluorescein succinimidyl ester (CFSE; Life Technologies). After the addition of serum and washes with PBS, cells were resuspended at 1 × 10^6^/mL and plated into 96-well U-bottom plates (Corning) at 200 uL volumes. Peptide pools were added at a final concentration of 0.25 ug/mL. On day 6, cells were harvested, washed with PBS + 2% Fetal Bovine Serum, and stained with anti-CD3-PE-Cy7 (clone SK7; BioLegend), anti-CD8 APC (clone SK1; BioLegend), anti-CD4 BV711 (clone RPA-T4; BioLegend) and LIVE/DEAD violet viability dye (Life Technologies). Cells were washed and fixed in 2% paraformaldehyde, prior to flow cytometric analysis on a BD LSR II (BD Biosciences). A positive response was defined as one with a percentage of CD3^+^ CD8^+^ or CD3^+^ CD4^+^ CFSE low cells at least 1.5x greater than the highest of two negative-control wells and greater than 0.2% CD8^+^ or CD4^+^ CFSE low cells in magnitude following background subtraction. For graphical analyses, responses are plotted at a value of 0.1% CD8^+^ or CD4^+^ CFSE low cells.

### Neutralization of wildtype and omicron spike pseudotyped virus

Neutralization data is from our recent study in a subset of individuals described here and previously reported (Garcia-Beltran et al., 2021b). In brief, pseudovirus neutralization titer 50 (pNT50) was calculated by taking the inverse of the serum concentration that achieved 50% neutralization of SARS-CoV-2 pseudotyped lentivirus particles entry into ACE2 expressing 293T cells (a gift from Michael Farzan). We introduced mutations corresponding to the SARS-CoV-2 variants of concern by site directed mutagenesis and confirmed clones by sequencing.

## QUANTIFICATION AND STATISTICAL ANALYSIS

The primary statistical analysis shown in Table S3 and S5 was a multivariate regression modelling T cell response (log_10_ CFU/10^6^ PBMC) as the response variable, and age, sex, peptide pool, prior infection, vaccine type, duration from vaccination as covariates. To compare proportions of individuals we used a fishers-exact test. Analyses were performed in R and figures rendered in GraphPad Prism. A p-value of < 0.05 was considered significant.

## SUPPLEMENTARY INFORMATION

## SUPPLEMENTARY FIGURE LEGENDS

**Figure S1: Effector T cell responses to spike and non-spike structural proteins are correlated for wildtype and Omicron amongst individuals with prior infection with or without vaccination, relates to Figure 1**. Shown are the correlations between magnitude of effector T cell response (IFN-g SFU per 10^6^ PBMCs) directed against spike and non-spike structural and accessory proteins (nucleocapsid, membrane, envelope and ORF3A) from wildtype (Panel A and C) or Omicron (B and D) spike. The upper panels A and B pertain to individuals with prior infection who were not vaccinated, and the lower panels depict prior infected and vaccinated individuals. In A-D each dot is a single participant. Circles denote responses to wildtype peptides and squares to Omicron peptides. Dots are colored by prior infection and vaccine stratum (blue prior infection and no vaccination; and orange for both prior infection and vaccination with primary series). Spearman correlation coefficients are denoted in each panel.

**Figure S2: Gating strategy for CFSE proliferation assay, relates to Figure 3**. Representative gating strategy for identification of proliferating CD3^+^ CD4^+^ and CD3^+^ CD8^+^ CFSE low T cells in response to peptide pools of interest. The gate establishing the frequency of CFSE low CD4 ^+^ or CD8 ^+^ cells was chosen based on minimizing responses in two negative-control (DMSO) wells and verified using positive control (CD3/CD28) wells.

## SUPPLEMENTARY TABLES

**Table S1: Baseline characteristics of included samples (N=101 from 76 unique donors), related to Figure 1**.

**Table S2: Details of variant peptide pools for spike and non-spike structural proteins, related to Figure 1**.

**Table S3: Multivariate regression of total T-cell response to SARS-CoV-2 spike peptides in vaccinated individuals, related to Figures 1 and 2**.

**Table S4. Frequency of individuals with greater than 50% (0.3log**_**10**_**) reduction in circulating effector T-cell response to variant, relative to wildtype, related to Figures 1 and 2**.

**Table S5: Multivariate regression of CD4+ and CD8+ T-cell proliferative response to SARS-CoV-2 spike peptides in vaccinated individuals, related to Figure 3**.

