## Supplemental Tables 1-5 for "T cell reactivity to the SARS-CoV-2 Omicron variant is preserved in most but not all prior infected and vaccinated individuals"

**Supplementary Appendix**

**Supplementary Tables**

**Table S1: Baseline characteristics of included samples (N=101 from 76 unique donors).**

|  | Unvaccinated, N = 21 | pre-boost, N = 36^#^ | post-boost, N = 44^#^ |
| --- | --- | --- | --- |
| **Age** | 34 (28, 58) | 46 (38, 54) | 48 (40, 64) |
| **Sex** |  |  |  |
| Female | 9 (43%) | 26 (72%) | 30 (68%) |
| Male | 12 (57%) | 10 (28%) | 14 (32%) |
| **Prior infection** |  |  |  |
| PCR/Antibody confirmed prior infection | 11 (52%) | 12 (33%) | 13 (30%) |
| No prior infection | 10 (48%) | 24 (67%) | 31 (70%) |
| **Vaccine type** |  |  |  |
| None | 21 (100%) | 0 (0%) | 0 (0%) |
| Ad26.COV2.S | NA | 4 (11%) | 7* (16%) |
| BNT-162b2 | NA | 24 (67%) | 27** (61%) |
| mRNA-1273 | NA | 8 (22%) | 10*** (23%) |
| **Duration after primary vaccination series (median days (IQR))** | NA | 220 (207, 243) | 232 (216, 250) |
| **Booster type** |  |  |  |
| heterologous | NA | NA | 7 (16%) |
| homologous | NA | NA | 37 (84%) |
| **Duration after booster (median days (IQR))** | NA | NA | 10 (9, 15) |
| # includes paired paired pre-boost and post-boost samples from 25 individuals | | |  |
| * includes 3 boosted with Ad26.COV2.S, 2 with mRNA-1273 and 2 with BNT162b2 | | | |
| ** includes 25 boosted with BNT162b2 and 2 with mRNA-1273 | |  |  |

**Table S2: Details of variant peptide pools for spike and non-spike structural proteins**

|  | **Spike** | | | | **Nucleocapsid, Membrane, Envelope, ORF3A** | |
| --- | --- | --- | --- | --- | --- | --- |
|  | **Wildtype** | **Delta** | **Omicron** | **Wildtype** | | **Omicron** |
| **Number of peptides in pool** | 315 | 315 | 315 | 237 | | 237 |
| **Peptides shared with wildtype** | Ref | 288 (91.4%) | 229 (72.7%) | Ref | | 213 (89.9%) |
| **Peptides unique to variant** | Ref | 27 (8.6%) | 86 (27.3%) | Ref | | 24 (10.1%) |

**Table S3: Multivariate regression of total T-cell response to SARS-CoV-2 spike peptides in vaccinated individuals.**

|  | **Effect estimate on total T-cell response (in log_10_ CFU/10^6^ PBMC)** | **95% Confidence Interval** | **p-value** |
| --- | --- | --- | --- |
| **Age (per 10 year increase)** | -0.03 | -0.09, 0.03 | 0.3 |
| **Sex** |  |  |  |
| Female | Ref |  |  |
| Male | -0.03 | -0.21, 0.15 | 0.7 |
| **SARS-CoV-2 variant** |  |  |  |
| ancestral | Ref |  |  |
| Delta (B.1.617.2) | -0.09 | -0.27, 0.09 | 0.3 |
| Omicron (B.1.1529) | -0.03 | -0.21, 0.15 | 0.7 |
| **Prior infection** |  |  |  |
| No prior infection | Ref |  |  |
| PCR/Antibody confirmed prior infection | 0.55 | 0.38, 0.72 | <0.001 |
| **Vaccine type** |  |  |  |
| mRNA-1273 | Ref |  |  |
| BNT-162b2 | 0.09 | -0.10, 0.28 | 0.3 |
| Ad26.COV2.S | -0.05 | -0.32, 0.22 | 0.7 |
| **Duration after primary vaccination series in weeks** | -0.02 | -0.05, 0.00 | 0.028 |
| **Booster group** |  |  |  |
| pre-boost | Ref |  |  |
| post-boost | 1.1 | 0.91, 1.2 | <0.001 |

**Table S4. Frequency of individuals with greater than 50% (0.3log_10_) reduction in in circulating effector T-cell response to variant, relative to wildtype**

|  |  | **Unvaccinated** | **pre-booster** | **post-booster** | **Overall** |
| --- | --- | --- | --- | --- | --- |
| **Omicron** | **Prior infected** | 27.3% (3/11) | 8.3% (1/12) | 15.4% (2/13) | 15.3% (14/91)* |
|  | **No prior infection** | - | 25% (6/24) | 6.5% (2/31) |  |
| **Delta** | **Prior infected** | 18.2% (2/11) | 0% (0/12) | 0% (0/13) | 5.9% (5/85) |
|  | **No prior infection** | - | 16.7% (3/18) | 0% (0/31) |  |
| *Fisher's exact test comparing frequency for Omicron vs. Delta in 85 individuals with both measured ie 13/85 5/85; p-value 0.024. | | | | | |

**Table S5: Multivariate regression of CD4+ and CD8+ T-cell proliferate response to SARS-CoV-2 spike peptides in vaccinated individuals.**

|  | | **CD4+ Memory T cell response** | | | | | | **CD8+ Memory T cell response** | | | | | |
| --- | --- | --- | --- | --- | --- | --- | --- | --- | --- | --- | --- | --- | --- |
|  | | **Effect estimate** | | **95% Confidence Interval** | | **p-value** | | **Effect estimate** | | **95% Confidence Interval** | | **p-value** | |
| **Age (per 10 year increase)** | | 0.16 | | -1.2, 1.5 | | 0.8 | | -0.67 | | -2.2, 0.88 | | 0.4 | |
| **Sex** | |  | |  | |  | |  | |  | |  | |
| Female | | Ref | |  | |  | |  | |  | |  | |
| Male | | 0.72 | | -3.1, 4.5 | | 0.7 | | 0.72 | | -3.1, 4.5 | | 0.7 | |
| **SARS-CoV-2 variant** | |  | |  | |  | |  | |  | |  | |
| ancestral | | Ref | |  | |  | |  | |  | |  | |
| Omicron (B.1.1529) | | -1 | | -4.5, 2.5 | | 0.6 | | -3.4 | | -7.4, 0.58 | | 0.093 | |
| **Prior infection** | |  | |  | |  | |  | |  | |  | |
| No prior infection | | Ref | |  | |  | |  | |  | |  | |
| PCR/Antibody confirmed prior infection | | 0.42 | | -3.2, 4.1 | | 0.8 | | 3.6 | | -0.54, 7.8 | | 0.087 | |
| **Vaccine type** | |  | |  | |  | |  | |  | |  | |
|  | mRNA-1273 | | Ref | |  | |  | | 4.1 | | -0.79, 9.0 | | 0.1 |
|  | BNT-162b2 | | 1.6 | | -2.7, 5.9 | | 0.5 | | 0.28 | | -9.0, 9.6 | | >0.9 |
|  | Ad26.COV2.S | | 2 | | -6.2, 10 | | 0.6 | |  | |  | |  |
| **Duration after primary vaccination series in weeks** | | -0.28 | | -0.74, 0.18 | | 0.2 | | -0.01 | | -0.53, 0.51 | | >0.9 | |
| **Booster group** | |  | |  | |  | |  | |  | |  | |
| pre-boost | | Ref | |  | |  | |  | |  | |  | |
| post-boost | | 3.2 | | -1.2, 7.5 | | 0.2 | | 5.4 | | 0.48, 10 | | 0.032 | |
