## Supplementary figures and images for "T cell reactivity to the SARS-CoV-2 Omicron variant is preserved in most but not all prior infected and vaccinated individuals"

### Supplemental FIgs 1-2

#### SUPPLEMENTARY FIGURE 1

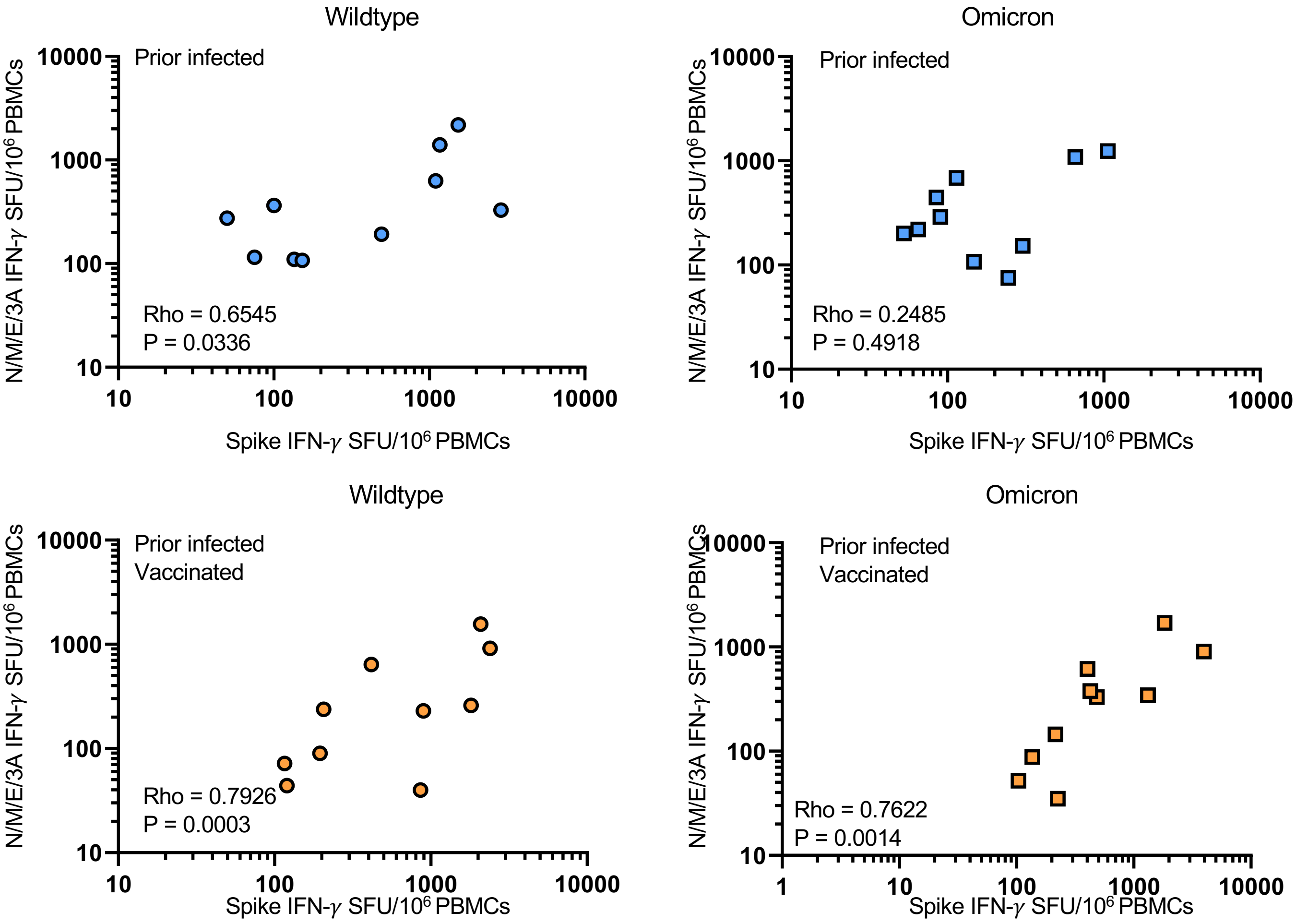

#### SUPPLEMENTARY FIGURE 2

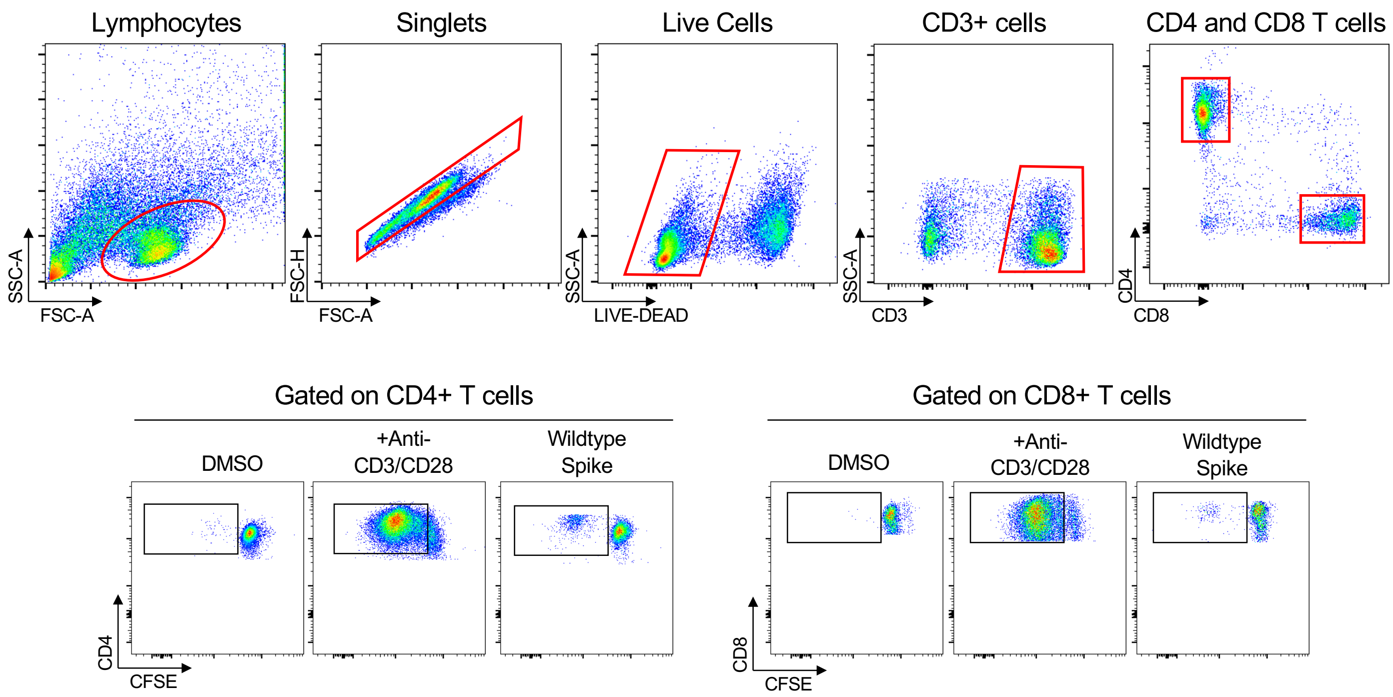
